# Caregivers and Coping: Well-being and coping styles among caregivers of young adults with developmental disabilities

**DOI:** 10.1101/2023.02.24.23286429

**Authors:** Hannah Singer, Elaine Clarke, Hillary Schiltz, Catherine Lord

## Abstract

Little research examines well-being in caregivers of individuals with developmental disabilities (DDs) during their child’s adulthood. Drawing from a longitudinal cohort of caregivers of adults with autism spectrum disorder (ASD) and other DDs (*n* = 134), this study examined change in caregiver well-being over ten years (young adults ages 18-28) and predictors of well-being, such as coping. Lower caregiver education and high young adult externalizing symptoms were related to lower caregiver well-being. Notably, caregiver coping style predicted degree of change in well-being over time. Effects of coping were moderated by demographic (i.e., parental education) and young adult characteristics (i.e., externalizing symptoms). In line with strengths-based approaches, these results illustrate the importance of coping styles in promoting caregiver well-being.

## Introduction

Autism spectrum disorder (ASD) and developmental disabilities (DD) are lifelong conditions. Concomitantly, for many parents, caregiving for children with ASD and/or DDs is a lifelong role This role encompasses both challenges (Maddox et al., 2017) and benefits (Marsack-Topolewski & Wilson, 2021) which can affect the psychological well-being of caregivers (Band-Winterstein & Avieli, 2017). Given that caregiver well-being may have cascading effects on family functioning (Dempsey & Keen, 2008), it is important to understand predictors of well-being among this population (Marsack-Topolewski & Church, 2019; Browne et al., 2015). Further, in order to consider caregivers’ strengths in addition to their challenges, improved understanding of potential protective factors, such as coping styles, is also needed. Existing literature points to varied predictors—including socioeconomic status (Lindsay & Barry, 2018), autism symptomatology (Smith et al., 2008), access to services (Harper et al., 2013), and caregivers’ coping styles (Wong et al., 2020)—that impact the well-being of caregivers. However, much of this research focuses on caregiver well-being during the childhood of the diagnosed individual. Additional work is needed to understand caregivers’ well-being as their children with autism and/or DDs enter adulthood.

Well-being likely evolves as caregivers age alongside their children. The first decade of adulthood (from late adolescence to late twenties) may be a particularly important period for understanding changes in caregiver well-being (Pellicano et al., 2022). This decade includes the transition out of school, sometimes termed the “services cliff” (Shattuck et al., 2012), when many individuals with autism and other DDs experience changes in (and often losses of) supports (McKenzie et al., 2017). Further, most studies of this population are cross-sectional, which limits understanding of temporal changes within individual caregivers, especially after key adulthood transitions (e.g., school exit).

Existing research also frequently relies on limited operationalizations of well-being. That is, many studies of caregivers of individuals with autism and/or DDs define well-being as an absence of negative constructs (i.e., stress, depression, anxiety; e.g., Herrema et al., 2017). Positively framed well-being, in contrast, includes constructs such as purpose in life, self-acceptance, and perceptions of personal growth (Deci et al., 2008; Ryff, 1989). Emphasizing these positive elements of well-being aligns with an on-going shift towards strength-based approaches throughout autism research (Urbanowicz et al., 2019). This type of strength-based approach highlights aptitudes and resiliencies in autistic and DD individuals and their families, alongside areas of need (Szatmari et al., 2021).

Consistent with a strengths-based perspective, coping can offer a constructive way to promote well-being in the face of stressors (Carver, 2013). Patterns of coping over time can be conceptualized as “coping styles” (Carver & Scheier, 1994) which can be behavioral or psychological (e.g., talking to a friend or reframing negative thoughts). They can also be productive or unproductive (e.g., meditating to relieve stress or punching a wall). Optimism and cognitive reframing-based coping have been linked to greater maternal ratings of happiness, enjoyment, and hopefulness about the future for caregivers of children with ASD (Wong et al., 2020; Benson, 2010). Research has also identified longitudinal effects of coping styles. For example, problem-focused coping may predict declines in depression among mothers of adults with intellectual disability (Kim et al., 2003). In contrast, increased distraction coping has been linked with increases in caregiver distress (Benson, 2014). Longitudinal approaches provide the opportunity to understand the implications of coping styles on individuals’ well-being as caregivers age and their children become adults. However, not all coping styles may be equally effective for all caregivers; research suggests that coping styles may differentially predict caregiver well-being based on characteristics of the individual with ASD (Benson, 2014). More work is needed to understand these longitudinal links in adulthood.

### Study Aims

Relatively little is known about well-being, especially from a strengths-based approach, among caregivers of adults with autism and/or DD. Our first aim was to characterize positive dimensions of well-being in caregivers over a ten-year period, when their young adults with autism and/or DDs were 18 to 28 years old. As part of our first aim, we also investigated effects of demographic (race, parental education, and child biological sex) and young adult person-level characteristics (ASD symptomatology, externalizing behaviors, and intellectual disability of the young adult) on change in caregiver well-being.

Although coping styles are a robust cross-sectional correlate of caregiver happiness, distress, and quality of life (Benson 2010; Benson 2014), limited work has examined longitudinal effects of coping styles on well-being, particularly in adulthood. To provide new information about caregivers’ resilience and inherent strengths, our second aim was to examine the impact of coping styles on patterns of caregiver well-being a ten-year period in our longitudinal study. As part of this second aim, we also examined whether the impact of coping styles on caregiver well-being was moderated by caregiver demographic and young adult personal characteristics.

## Method

### Participants

The current sample draws from a longitudinal study of 253 individuals referred for autism evaluations and their families. Data for this paper was collected over ten years, when the young adults with autism and/or DDs were ages 18 to 28. Given this study’s focus on caregiver well-being, these analyses included 134 caregivers (*M*age=48.1, SD=6.04) who completed the Scales of Psychological Well-Being questionnaire (Ryff, 1989) at least once when their young adult was between ages 18-28. Twenty-nine caregivers had young adults with diagnoses of a DD other than ASD; 105 had diagnoses of ASD. Among the sample’s caregiver respondents, there were 118 mothers, 11 fathers, 3 grandmothers, and 2 aunts. A majority of caregivers were white (82%), non-Hispanic (98%), completed a college degree (65%), and had a male young adult (79%). Differences between participants in the current subsample and the total longitudinal cohort are reported in Table 1. Chi-square analyses revealed race and caregiver education significantly differed between the current subsample and total sample, with fewer participants of color and fewer caregivers with some college or less in our subsample. Race and caregiver education were therefore treated as covariates in all analyses.

**Table 1.** Demographic Characteristics of the Current Subsample and Total LSA sample.

|  |  | **Study Subsample**<br>**(N=134)** | **Total LSA**<br>**Sample (N=253)** |  |
| --- | --- | --- | --- | --- |
| **Race** | Black or Asian | 25 | 74 | **$X^2(1) = 15.446;$**<br>**$p = <.001$** |
|  | White | 109 | 179 |  |
| **Child**<br>**Biological Sex** | Male | 106 | 203 | $X^2(1) = .230;$<br>$p = .631$ |
|  | Female | 28 | 50 |  |
| **Caregiver**<br>**Education** | Some college or less | 46 | 114 | **$X^2(1) = 48.894;$**<br>**$p = <.001$** |
|  | College degree or more | 88 | 139 |  |
| **ASD Status** | ASD | 105 | 196 | $X^2(1) = .129;$<br>$p = .720$ |
|  | Other DD | 29 | 57 |  |
| **Urbanicity** | Rural | 42 | 88 | $X^2(1) = 1.220;$<br>$p = .269$ |
|  | Urban | 87 | 158 |  |

### Procedure

All studies were approved by relevant IRBs; guardians and individuals with ASD and/or DDs over 18 who were their own legal guardians gave written consent as required by the relevant institutional review board(s) (IRB) prior to visits. Questionnaires were mailed to caregivers six times, when young adults with ASD and/or DDs were approximately 18, 20, 22, 24, 26, and 28 years old. When young adults with ASD and/or DDs were approximately age 18, face-to-face testing, including the Autism Diagnostic Interview-Revised (ADI-R, Le Couteur et al., 2003), Autism Diagnostic Observation Schedule, Second Edition (ADOS-2; Lord et al., 2012), the Vineland Adaptive Behavior Scales, Second Edition (VABS-2; Sparrow et al., 1984, 2005), and standardized developmentally appropriate IQ tests (see Anderson et al., 2014) were conducted by clinicians blinded to diagnosis and previous test results.

### Measures

#### Well-being

Caregivers completed the Scales of Psychological Well-Being (SPWB, by Ryff, 1989) multiple times throughout the study (*M* = 3.88, range = 1-6). The SPWB has well-documented reliability and validity across age groups, and has been used across a variety of contexts, including longitudinal studies (Abbott et al., 2010). Consistent with SPWB author recommendations to reduce participant burden (Ryff, 1989), caregivers (n=134) completed an abbreviated version of the SPWB containing 48 items rated on a 6-point Likert scale. The abbreviated SPWB subscales included the following: self-acceptance (e.g. items like, “In general, I feel confident and positive about myself”), personal growth (e.g., “I enjoy seeing how my views have changed and matured over the years”), and purpose in life (e.g., “I feel good when I think of what I’ve done in the past and what I hope to do in the future”). Subscales were summed to calculate total well-being scores.

#### Coping

Caregivers (*n* = 99) completed the Brief Coping Orientation to Problems Experienced Inventory (Brief-COPE; Carver et al., 1997) when their young adults were approximately 18 years old. The Brief-COPE is a well-validated and reliable measure, and offers several strengths, including ease of completion and measurement of a range of coping responses (Muniandy et al., 2021). The Brief-COPE consists of 28 items rated on a 4-point Likert scale that describe strategies used in response to stressors (e.g., “I’ve been saying to myself ‘this isn’t real’”). In the current study, caregivers were specifically instructed, “These items ask what you’ve been doing to cope with the challenges you may encounter raising a child on the autism spectrum.”

Brief-COPE subscales were based on empirically-derived factors from a prior study of caregivers of children with ASD (Hastings et al.; 2005). For the sake of interpretability and clarity, the “religious/denial coping” subscale was scored as two separate subscales (i.e., **religious** and **denial**), producing a total of five coping dimensions: **active avoidance** (attempts to “avoid” the stressor); **problem-focused** (attempts to directly confront the stressor to modify or eliminate its effects); **positive** (use of humor, positive reframing, acceptance, and emotional social support); **religious** (praying, meditation, or seeking comfort in religion), and **denial** (attempts to tell oneself the situation is not real).

#### Externalizing Behaviors

Caregivers completed the Child Behavior Checklist (CBCL; Achenbach & Rescorla, 2001) when their young adults were approximately 18 (n = 111). Containing 118 items on a 3-point Likert scale, the CBCL has evidence for validity among individuals with ASD (Schiltz & Magnus, 2020). Based on previous studies that found externalizing behaviors predict caregiver outcomes (Rattaz et al., 2017; Wong et al., 2020), the current study used externalizing domain scores, which include CBCL’s rule-breaking behavior and aggressive behavior subscales.

#### IQ

Cognitive abilities of the adults with ASD and/or DDs were measured at approximately age 18 via face-to-face assessments selected from a standardized hierarchy (n=117). Ratio IQs were calculated from age equivalents when raw scores fell outside deviation score ranges. See Anderson et al., 2014 for a detailed description of IQ testing in this sample.

#### Diagnoses and Autism Symptomology

Clinicians made blinded diagnoses of ASD, other neurodevelopmental disabilities, or typical development at each in-person visit. Twenty-nine individuals in this sample had diagnoses of DDs other than autism, and 105 individuals had autism. Given considerable overlap in challenges faced during young adulthood for all individuals with developmental disabilities and their caregivers (Lord et al., 2022), all participants with or without DD, were retained in the current analyses.

Autism symptomology of every young adult, regardless of diagnosis, was measured using calibrated severity scores (CSS) from the Autism Diagnostic Observation Schedule, Second Edition (ADOS-2; Lord et al., 2012) at approximately age 18 years old (n = 107), and when unavailable, from the next closest assessment. CSS scores range from 1 to 10, with higher scores reflecting more autism symptoms.

#### Daily Living Skills

Clinicians administered the Vineland Adaptive Behavior Scales, 2nd edition (VABS-II, Sparrow et al., 2005) to caregivers when their adults with DDs were approximately 18 years old. The VABS-II is a standardized and well-validated structured interview assessing communication, daily living skills (DLS), socialization, in comparison to other same-aged individuals. DLS age equivalence (AE) scores were used in the current analyses.

### Data Analysis

#### Multilevel Modeling

Multilevel modeling via the mixed procedure in Stata 17.0 was used to investigate changes in caregiver well-being over the 10-year study period, as well as the impact of demographic characteristics and coping styles on the intercept of parent well-being and changes in well-being over time (i.e., cross level interactions with age). Time (i.e., age) was centered at the adult child’s age 18 for ease of interpretation. A multilevel modeling approach was selected to account for the nested structure of data across time and non-independence of assessment observations, and to evaluate the inclusion of random effects across participants on well-being estimates. For our multilevel models, we used an alpha level of .05 for null hypothesis significance testing.

##### Aim 1: Impacts of Demographic and Child Characteristics on Well-Being

First, the effects of demographic covariates (i.e., caregiver education, race) and young adult-level factors (i.e., IQ, externalizing behavior, daily living skills) on caregiver well-being were modeled independent of coping styles. Effects of demographic and young adult-level factors were tested as main effects and as interactions with time.

##### Aim 2: Associations Between Coping Styles and Well-Being

Models examining the effect of each coping style on caregiver well-being were constructed and analyzed separately. This approach was used for several reasons. First, by examining each coping style seperately, we aimed to gain a more fine-grained understanding of the independent effects of each coping style on caregiver well-being. This approach also allowed us to avoid issues with multicollinearity. Models were constructed using a three-step approach. The first and simplest model for each coping style included fixed effects for time, coping style, time x coping style interaction terms, and a random effect for time. Next, demographic covariates and young adult factors (e.g., race, recruitment site, caregiver education, participant IQ, externalizing behavior, CSS, and DLS-AE) were added to models as fixed effects. Non-significant effects were then removed. Finally, for significant covariates, covariate x coping style, covariate x time, and covariate x coping style x time interaction terms were added. Non-significant covariates and interaction terms identified at this stage were removed from the final models.

## Results

### Effects of Caregiver Demographic and Adult Child Factors on Well-Being

The first model examined change in parent well-being, without accounting for variance explained by coping style (Table 2). There was no significant fixed effect of time on caregiver well-being, indicating stability in well-being on average across the ten years. Random effects for participant and time were significant (both *p* > .05), indicating average well-being scores differed across individual caregivers and the change in well-being over time differed across individual caregivers, respectively. There were no significant effects of race, child IQ, DLS, ADOS CSS, urbanicity, child biological sex, and recruitment site (all *p* > .05). These covariates were dropped from the final well-being model. There were significant main effects of caregiver education (b = 16.612, *p* = .001), and child externalizing symptoms (b = -.683, *p* = .002). On average, caregivers with less than a college degree and caregivers of individuals with high externalizing symptoms had lower well-being across the study before we accounted for caregivers’ use of various coping styles. Interaction terms between significant covariates by time were tested, but none were significant. In other words, not considering coping, the degree of change in caregiver well-being over time was not impacted by demographic factors.

**Table 2.** Demographic Characteristics of the Current Subsample and Total LSA sample.

| Caregiver Total Well-being on SPWB |  |  |  |  |  |
| --- | --- | --- | --- | --- | --- |
| Main Effects |  | Coefficient | SE | 95% CI |  |
| Caregiver education |  | 16.612** | 4.625 | 7.545 | 25.678 |
| Race |  | -- | -- | -- | -- |
| Urbanicity |  | -- | -- | -- | -- |
| Child Externalizing (CBCL) |  | -0.683** | 0.223 | -1.121 | -0.245 |
| Child Biological Sex |  | -- | -- | -- | -- |
| Child Non-Verbal IQ |  | -- | -- | -- | -- |
| Child Autism Symptoms (CSS) |  | -- | -- | -- | -- |
| Child Daily Living Skills (VABS-2) |  | -- | -- | -- |  |
| Time |  | 0.231 | 0.299 | -0.356 | 0.818 |
| Random Effects |  | Coefficient | SE | 95% CI |  |
| Time |  | 29.509 | 17.614 | 9.159 | 95.071 |
| Individual Caregiver |  | 460.504 | 73.607 | 336.648 | 629.929 |

*Note.* Table 2 displays multilevel output of impacts of demographic and child characteristics on trajectories of caregiver well-being. Non-significant covariate main effects were identified and omitted before testing interaction terms. Non-significant covariate interaction terms were removed from final well-being models.
\*\*Significant at .01.

### Effects of Caregiver Coping Styles on Well-Being

#### Positive Coping

Main effects of positive coping (b = 7.785, and time (b = -1.953, *p* = .121) on caregiver well-being were not significant, nor was the interaction between time x positive coping (b = -1.045, *p =* .152; Table 3). There was, however, a significant three-way externalizing x positive coping x time interaction (b = .039, *p* = .0001). The effect of positive coping on change in well-being differed based on externalizing behaviors of the young adult with autism and/or DD. Positive coping appeared more beneficial (i.e., related to larger, positive differences in well-being) for caregivers of individuals with high (vs lower) levels of externalizing symptoms at age 18 (Figure 1).

**Table 3.** Effects of caregiver coping styles on caregiver well-being over time.

|  | Coping Style |  |  |  |  |  |  |  |  |  |  |  |
| --- | --- | --- | --- | --- | --- | --- | --- | --- | --- | --- | --- | --- |
|  | Positive Coping |  |  |  | Active Avoidance Coping |  |  |  | Problem-Focused Coping |  |  |  |
|  | Coefficient | SE | 95% CI |  | Coefficient | SE | 95% CI |  | Coefficient | SE | 95% CI |  |
| Coping Style | 7.785 | 4.083 | -.218 | 15.789 | -12.804 | 6.990 | -26.505 | .896 | -47.608** | 18.288 | -83.453 | -11.764 |
| Caregiver Education | -- | -- | -- |  | 14.522** | 5.022 | 4.679 | 24.366 | -- | -- | -- |  |
| Race | -11.658 | 6.082 | -23.580 | .262 | -- | -- | -- |  | -64.775** | 24.063 | -111.938 | -17.612 |
| CBCL Externalizing | -1.233** | .250 | -1.725 | -.742 | -.781** | .249 | -1.271 | -.292 | -3.047** | .930 | -4.871 | -1.223 |
| Time | -1.953 | 1.258 | -4.419 | .513 | -2.824* | 1.127 | -5.034 | -.613 | -4.659 | 2.451 | -9.463 | .144 |
| Coping Style X Time | -1.045 | 0.728 | -2.473 | .383 | 2.097** | .735 | .656 | 3.539 | 2.369* | 1.077 | .257 | 4.481 |
| Race X Time | -- | -- | -- |  | -- | -- | -- |  | 7.834** | 2.785 | 2.375 | 13.293 |
| Race X Coping Style | -- | -- | -- |  | -- | -- | -- |  | 22.544* | 9.514 | 3.897 | 41.191 |
| CBCL Externalizing X Coping Style | -- | -- | -- |  | -- | -- | -- |  | .788* | .343 | .115 | 1.462 |
| Race X Time X Coping Style | -- | -- | -- |  | -- | -- | -- |  | -3.479** | 1.180 | -5.793 | -1.165 |
| CBCL Externalizing X Time X Coping Style | .039** | .011 | .017 | .060 | -- | -- | -- |  | -- | -- | -- |  |
|  | Positive Coping |  |  |  | Active Avoidance Coping |  |  |  | Problem-Focused Coping |  |  |  |
|  | Coefficient | SE | 95% CI |  | Coefficient | SE | 95% CI |  | Coefficient | SE | 95% CI |  |
| Time | 27.493* | 17.369 | 7.970 | 94.842 | 35.219* | 19.984 | 11.582 | 107.096 | 27.912* | 18.102 | 7.829 | 99.501 |
| Individual Caregiver | 455.876* | 78.190 | 325.726 | 638.031 | 457.654* | 79.010 | 326.275 | 641.935 | 427.826* | 74.009 | 304.801 | 600.505 |

Table 3 (continued)
|  | Coping Style |  |  |  |  |  |  |  |
| --- | --- | --- | --- | --- | --- | --- | --- | --- |
|  | Religious Coping |  |  |  | Denial Coping |  |  |  |
|  | Coefficient | SE | 95% CI |  | Coefficient | SE | 95% CI |  |
| Coping Style | 4.857* | 2.430 | 0.938 | 9.620 | -10.815 | 7.071 | -24.675 | 3.045 |
| Caregiver Education | 14.519** | 4.991 | 4.736 | 24.302 | 14.738** | 4.965 | 5.007 | 24.470 |
| Race | -- | -- | -- | -- | -- | -- | -- | -- |
| CBCL Externalizing | -.774** | .241 | -1.248 | -.301 | -.780** | .244 | -1.260 | -.300 |
| Time | 2.697** | .740 | 1.246 | 4.148 | -2.456** | .860 | -4.142 | -.769 |
| Coping Style X Time | -.991** | .273 | -1.526 | -.455 | 2.348** | .699 | .978 | 3.719 |
| Race X Time | -- | -- | -- | -- | -- | -- | -- | -- |
| Race X Coping Style | -- | -- | -- | -- | -- | -- | -- | -- |
| CBCL Externalizing X Coping Style | -- | -- | -- | -- | -- | -- | -- | -- |
| Race X Time X Coping Style | -- | -- | -- | -- | -- | -- | -- | -- |
| CBCL Externalizing X Time X Coping Style | -- | -- | -- | -- | -- | -- | -- | -- |
|  | Religious Coping |  |  |  | Denial Coping |  |  |  |
|  | Coefficient | SE | 95% CI |  | Coefficient | SE | 95% CI |  |
| Time | 21.430* | 16.826 | 4.599 | 99.856 | 21.496* | 17.692 | 4.283 | 107.879 |
| Individual Caregiver | 471.419* | 80.112 | 337.874 | 657.748 | 462.274* | 78.966 | 330.747 | 646.104 |

*Note.* For length, only covariates with significant main effects and/or interaction terms in at least one coping model are reported in Table 3. For all coping models, non-significant main effects were identified and omitted before testing interaction terms, and non-significant interaction terms were removed from final coping models.
\*Significant at .05. \*\*Significant at .01.

**Figure 1.**
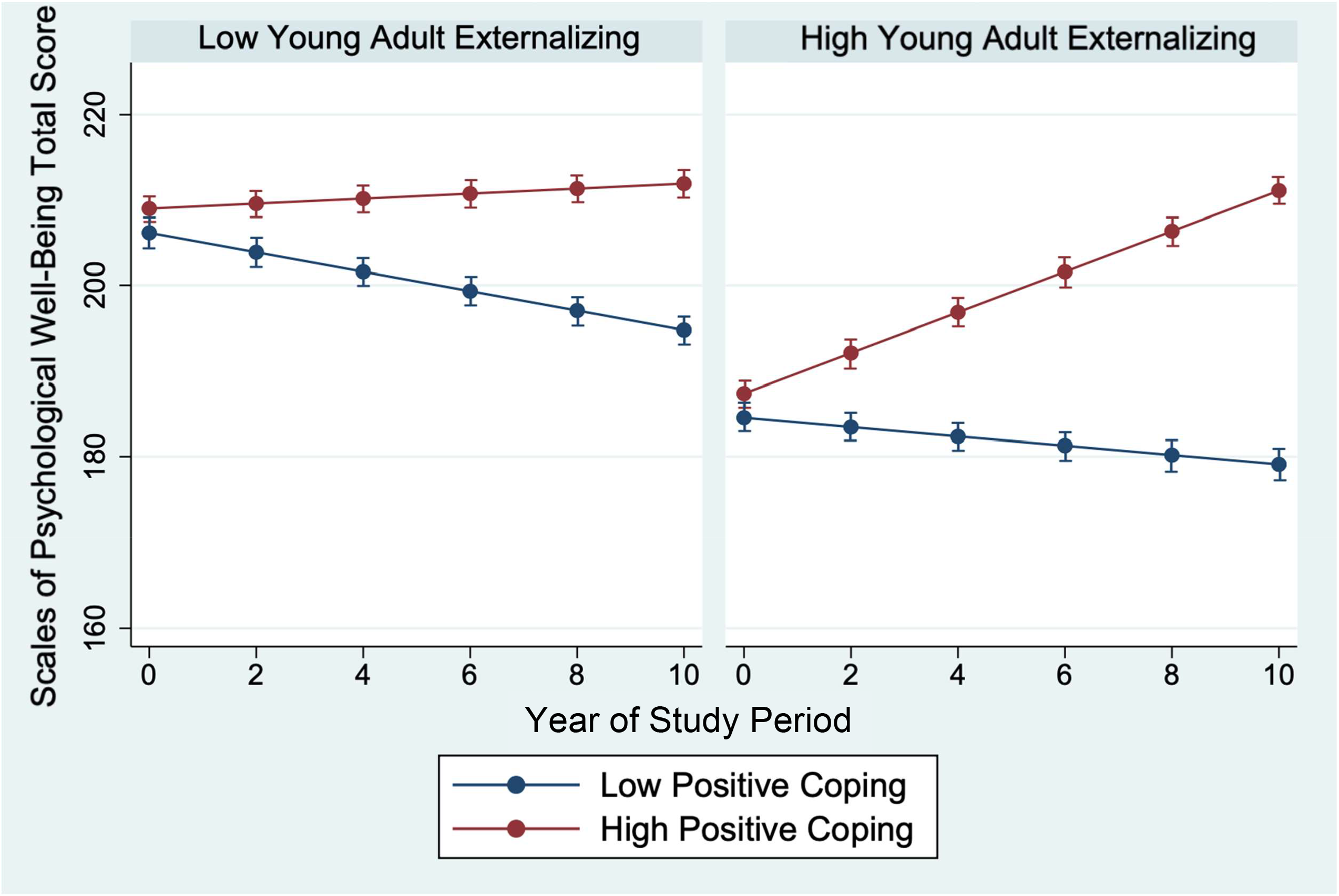
Effect of Positive Coping on Caregiver Well-Being by Young Adult Externalizing *Note*. For all coping style models, significant time x covariate interactions were probed by plotting well-being across time for all possible combinations of dichotomized covariates. To graph significant interactions with continuous predictors (e.g., CBCL externalizing scores), we created dichotomized variables, in which 0 indicated that an individual’s score for that continuous predictor was below the sample mean and 1 indicated an individual’s score was equal to or greater than the sample mean.

#### Active Avoidance Coping

The main effect of active avoidance coping was not significant (b = -12.804, *p* = .067). However, there was a significant two-way interaction between active avoidance coping x time (b = 2.09, *p* = .004). Caregivers who reported higher active avoidance coping at the beginning of the study demonstrated increasing well-being over time, while caregivers who reported lower active avoidance coping demonstrated consistent well-being over time.

#### Problem-Focused Coping

There was a main effect of problem-focused coping (b = -47.608, *p* = .009) and a significant time x problem-focused coping interaction (b = 2.369, *p* = .028) on caregiver well-being. There was also a significant two-way interaction between child externalizing x problem-focused coping (b = .788, *p* = .022; Figure 2) and a significant three-way interaction between race x problem-focused coping x time (b = -3.479, *p* = .003; Table 3; Figure 3). Problem-focused coping predicted well-being more for caregivers of adults with high externalizing symptoms than those with low externalizing symptoms, particularly early in the study (Figure 2). Further, the effect of problem-focused coping on parent well-being during the study period differed by caregiver race (Figure 3). Caregivers of color demonstrated an effect of problem focused coping on the change in well-being over time, while no such effect was evident for white caregivers.

**Figure 2.**
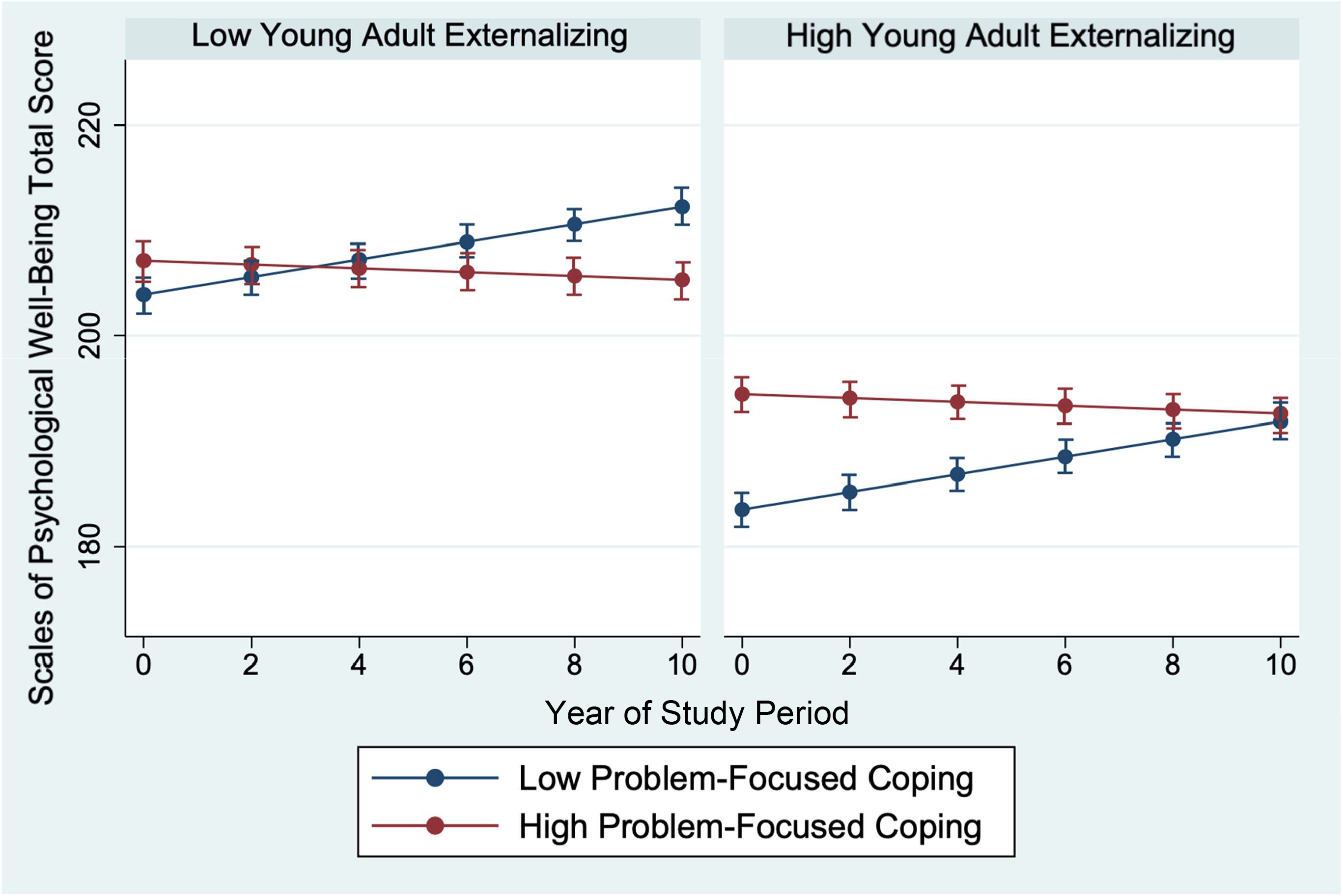
Effect of Problem-Focused Coping on Caregiver Well-Being by Young Adult Externalizing

**Figure 3.**
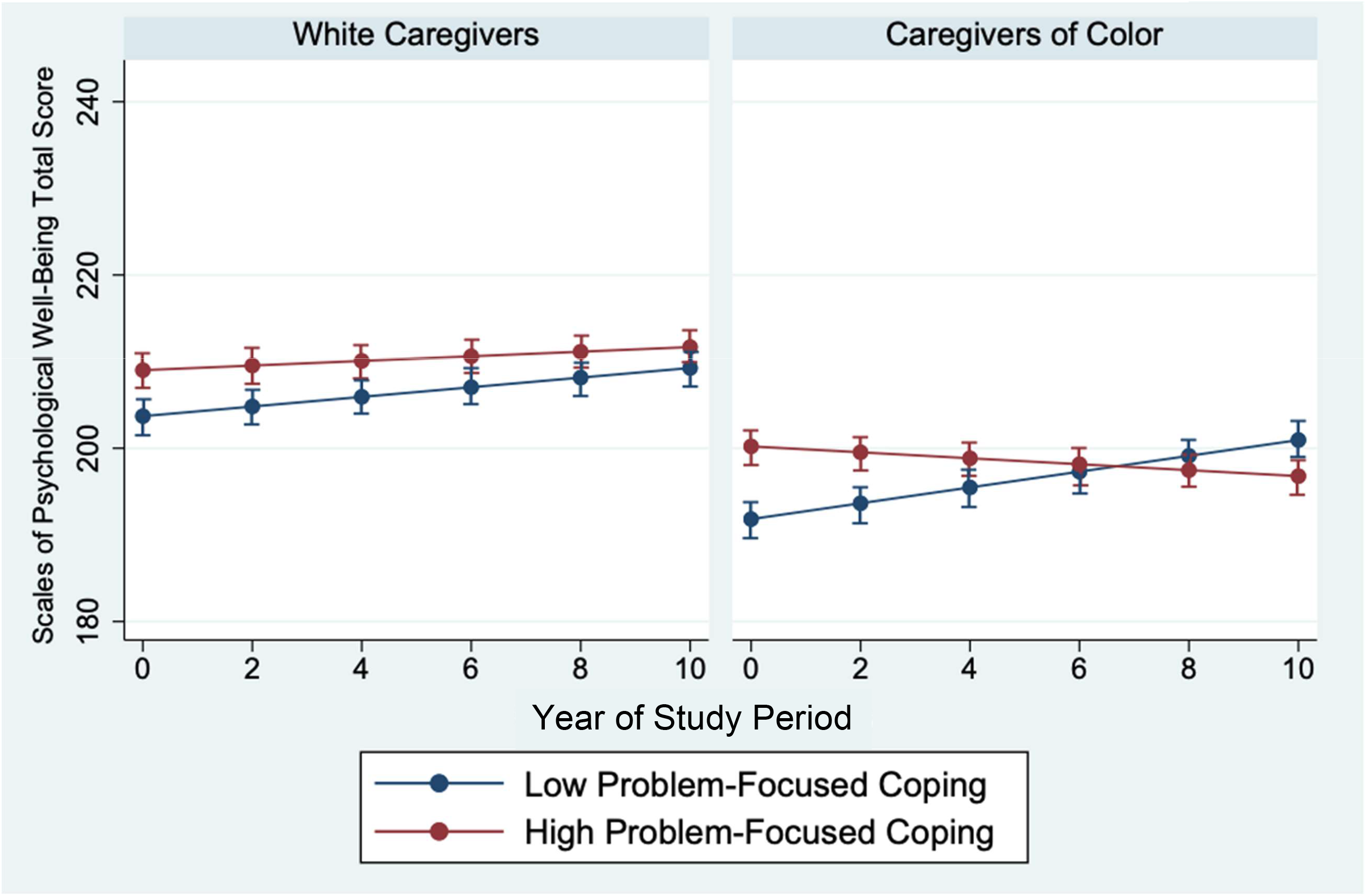
Effect of Problem-Focused Coping on Caregiver Well-Being by Caregiver Race

#### Religious Coping

There was a significant main effect of religious coping (b = 4.857, *p =* .046) on caregiver well-being and a significant religious coping x time interaction (b = -.991, *p <* .001; Table 4). Caregivers who reported low religious coping at the beginning of the study had lower well-being than those who reported high religious coping, but by the end of the study period, low religious copers reported greater well-being than those who used more religious coping.

#### Denial Coping

There was no main effect of denial coping (b = -10.815, *p* = .126), but there was a significant interaction between time and denial coping (b = 2.348, *p* = .001). Caregivers who engaged in denial coping reported increasing well-being over time. Caregivers who reported low denial coping, on the other hand, reported consistent well-being over time.

## Discussion

Caregiver well-being is a critical aspect of family functioning (Browne et al., 2015). However, despite the many challenges associated with the transition to adulthood for individuals with autism and/or DDs, little is known about stability and change in the well-being of their caregivers during this period. All too often, the literature on caregivers of individuals with autism and DDs emphasizes negative outcomes and detrimental aspects of caring for those with disabilities. The current study used a strengths-based approach—employing a positive operationalization of well-being and examining styles of coping with caregiving experiences—to address this literature gap. By examining the unique effects of both adaptive and maladaptive caregiver coping styles on positively-framed well-being, we sought to highlight the resilience of this population, and convey the experiences of these caregivers complexly and with dignity. We feel it is imperative to capture their strengths alongside their struggles.

To accomplish this, we first examined change in caregiver well-being over a ten-year period (when caregivers’ young adults were 18 to 28). Second, we identified demographic factors (e.g., caregiver education) and coping styles (e.g., use of positive coping) that significantly predicted change in caregiver well-being over time. Altogether, systematic variations in caregiver well-being were only apparent once the unique effects of coping styles on change in well-being were considered.

### Influences of Demographic and Adult Child Factors on Caregiver Well-Being

Only caregiver education and young adult externalizing symptoms predicted caregiver well-being during their child’s adulthood, independent of coping styles. On average, caregivers in our sample with less than a college degree reported lower well-being. Lower caregiver education is often associated with greater economic insecurity (Hill & King, 1995), and such economic stressors likely take a toll on caregivers (Conger & Conger, 2002; Gard et al., 2020). For example, financial hardship can exacerbate parenting stress (Trentacosta et al., 2018) and structural barriers (e.g., transportation, costs) to meeting child and family needs (Pickard & Ingersoll, 2016). Caregivers of young adults with lower levels of externalizing behaviors reported higher caregiver well-being across the study period. Such results suggest the effects of externalizing or “challenging” behaviors (Rattaz et al., 2017; Wong et al., 2020), may have a more pronounced impact on caregivers’ well-being than other individual differences such as the adult’s cognitive ability or self-care needs. This is consistent with previous findings that general behavior difficulties (Schiltz & Van Hecke, 2020; Wong et al., 2020), rather than ASD-specific symptoms (Benson, 2006), may drive lower caregiver well-being in this population.

Notably, young adult autism symptoms, IQ, and daily living skills did not predict well-being, and there were no significant differences in well-being between caregivers of adults with DDs and caregivers of adults with autism spectrum disorder with or without intellectual disability. The null effects of these young adult characteristics are striking given the range of symptoms and abilities in our sample. Prior work in this sample (Lord et al., 2020; Clarke et al., 2021) and others (Mawhood, Howlin, & Rutter, 2000; Zwicker, Zaresani, & Emery, 2017) has found adults with ASD and DDs experience similar difficulties attaining normative outcomes, such as employment and living independently. Relatedly, caregivers of children with ASD and DDs report comparable experiences of stigma (Mitter, Ali, & Scior, 2019) and mental health challenges (citation). This study adds to a growing body of literature finding more similarities than differences in the experiences of adults with autism and DDs, as well as their families.

### Influences of Coping Styles on Caregiver Well-Being

This study examined the impact of five coping styles (positive, active avoidance, problem-focused, religious, and denial) on caregiver well-being trajectories, and whether coping styles interacted with demographic and adult child characteristics. We found all five coping styles were significantly associated with change in caregiver well-being. Positive coping predicted higher caregiver well-being over time, especially for caregivers whose adult children exhibited high externalizing behaviors. The same trend was found for problem-focused coping. However, though problem-focused coping was associated with higher well-being intercepts at the first timepoint for caregivers of color and caregivers of high externalizing adults, it had less of an impact on well-being over time. In other words, caregivers with low problem-focused coping demonstrated similar levels of well-being to those with high problem-focused coping by the end of the ten-year study period. Individuals with neurodevelopmental disorders and their families lose access to school-based services and other potential sources of instrumental support in the years surrounding the transition to adulthood (Laxman et al., 2019; Shattuck et al., 2011). This loss of services is associated with declines in daily living skills (Clarke et al., 2020) and slowing improvements in social communication (Taylor & Seltzer, 2010). Thus, for caregivers who frequently engage in problem-focused coping, the dwindling availability of supports and lack of adult services may make the first decade of adulthood an especially frustrating time. Our results indicate this may be especially true for caregivers of color and caregivers of adults with challenging behaviors; additional work in separate samples is needed to replicate these findings.

Counterintuitively, we also found both denial coping and active avoidance coping were related to improved caregiver well-being over time. Although these coping strategies are typically considered “maladaptive,” perhaps they offer some benefit for caregivers of adults with DDs. Given the complexities of transitioning to adulthood that affect both adults with ASD and DDs and their caregivers, it may be helpful to create some mental distance between sometimes harsh realities and caregivers’ internal experiences by avoiding or denying their adult children’s ongoing challenges, choosing to view their circumstances with a rosier picture. In contrast, use of religious coping was associated with higher caregiver well-being at the beginning of the study period, but declines in well-being over time. There is some evidence that informal sources of support, such as friends or church groups, become decreasingly available or helpful for caregivers of individuals with ASDs over time (Anderson et al., 2018). Perhaps this declining availability may be reflected in the decreasing well-being of caregivers in our sample who endorsed using religious coping.

Coping styles did not interact with caregiver education. Given this and prior findings regarding adults with lower education and SES reporting lower well-being overall, coping styles may not buffer the circumstances faced by caregivers in lower socioeconomic groups. Interestingly, prior work in this sample found caregivers of color (Bishop et al., 2007) and caregivers with less than a college degree (Carr & Lord, 2013) report less perceived burden from caring for their child with ASD and/or DDs than white and college-educated caregivers. Well-being and perceived burden are related but distinct constructs. Further work is needed to fully characterize well-being and burden associations in caregivers of individuals with ASD or DDs.

### Clinical Implications

Intervention literature on caregivers of individuals with autism often focuses on supporting caregivers immediately following diagnosis (Catalano et al., 2018). However, these interventions are not designed to support caregiver well-being over many years (Prata et al., 2018). Considering the high rates of anxiety and depression among caregivers of people with intellectual disabilities (Gogoi et al., 2017) and with autism (Scherer et al., 2019), there is a clear need to support caregiver’s psychological functioning as their child ages. Our results suggest teaching and supporting coping strategies may be a promising route to promote caregiver well-being among this population. Given that positive coping predicted greater well-being, especially for caregivers of adults with high externalizing behaviors, interventions leveraging aspects of positive coping might be useful for this population. For example, Acceptance and Commitment Therapy (ACT), which emphasizes accepting difficult thoughts and emotions, has been found to be helpful for caregivers of children with ASD (Blackledge & Hayes, 2006; Corti et al., 2018) and therefore may be a fruitful therapeutic approach.

### Limitations

Despite the strengths of this study, including the longitudinal design, this work is not without limitations. In particular, although theory suggests coping styles are relatively stable (Carver & Scheier, 1994), this study measured caregiver coping styles at only one timepoint. Therefore, we were unable to account for potential change and/or stability in coping styles over time in our analyses. Further, a unique and relatively small group of families comprise this sample. Initially evaluated in the early 1990s, these caregivers sought help early in childhood, during an era in which knowledge about autism and DDs was less widespread than today. This sample’s participants may therefore differ meaningfully from individuals diagnosed in more recent decades and/or later in development. Finally, though the current sample remains more racially and educationally representative than many other samples of autism (Steinbrenner et al., 2022), the participation rates of Black families and families with lower caregiver education have decreased over the study’s 30-year-course.

### Future Directions

By focusing on the benefits of coping and positive elements of well-being, the current analyses leveraged a strengths-based approach to understanding outcomes for caregivers of adults with autism and other DDs. There have been recent calls for an increase in strengths-based empirical investigations into individuals with ASD (Szatmari et al., 2021); we feel strongly that more strengths-based studies at the family systems level are needed as well. There is an ample body of work detailing the negative effects of caregiving for a family member with an DD, including increased symptoms of depression (Scherer, Verhey, & Kuper, 2019), increased healthcare costs (Buescher et al., 2014) and decreased family income (Cidav et al., 2012). Yet many caregivers show lifelong resilience in the face of these chronic challenges. Families often report caring for an individual with an DD is a source of great meaning and joy—as well as stress. Future work should strive to capture these complexities and to elevate and support caregivers’ inherent capacities for resilience.

We identified some counterintuitive associations between coping styles and caregiver well-being across the ten-year study period. Two maladaptive coping styles, active avoidance and denial, were associated with increased well-being over time, while religious coping was associated with decreases in well-being. Future work should attempt to replicate and further examine these associations.

Finally, coping styles present a potential target for tailored intervention. Future studies of interventions for caregivers of adults with DDs could teach and promote the use of certain coping styles tailored to family and child characteristics, for example, prompting positive coping strategies amongst caregivers of individuals with externalizing behaviors.

## Conclusion

The life course impacts for those with DDs and autism are felt not only by diagnosed individuals, but by their caregivers and families. This study characterizes stability and change in the well-being of caregivers of young adults with autism and other DDs, and sheds light on the interactive effects of coping styles, family demographic characteristics, and young adult characteristics on caregiver well-being during this unique developmental period. Caregivers with less than a college degree reported lower well-being. Caregivers of adults with autism and/or DD with greater externalizing behaviors also reported lower well-being, but those who used positive and problem-focused coping appeared to benefit. Active avoidance and denial coping were associated with increased well-being over time; in contrast, religious coping was associated with declines in caregiver well-being. As caregivers and their young adults with ASD or DDs age and formal channels of support become increasingly limited, identifying internal resources that support caregiver well-being becomes increasingly pivotal. Coping styles may be leveraged to help caregivers in the face of difficult circumstances for their families.

## Data Availability

All data produced in the present study are available upon reasonable request to the authors

